# Should We Keep Changing the Clock? Characterizing Causal Effects of Daylight Saving Time on Behavior and Physiology

**DOI:** 10.64898/2025.12.20.25342749

**Authors:** Hayoung Jeong, Srikar Katta, Will Ke Wang, Alexander Volfovsky, Jessilyn Dunn

## Abstract

Daylight Saving Time (DST) remains contentious: some policymakers highlight behavioral benefits, while others emphasize health risks. Existing evidence relies largely on aggregated data and strict modeling assumptions, obscuring individual-level patterns and yielding associational rather than causal insights. We address these limitations using objective, longitudinal Fitbit measures from the NIH *All of Us* Research Program. Avoiding strict modeling assumptions, we employed a natural difference-in-differences design using Arizona (no DST) as a control against neighboring Mountain Time states (observing DST). Contrary to the common belief, DST transitions produced no net change in total daily steps. Instead, activity was reallocated to other times of day: fall transitions increased morning steps by 202 while reducing evening steps by 180; spring showed the opposite. Importantly, these treatment effects varied by demographics: older adults (65+) and lower-income individuals showed greater morning reallocation during fall DST, while younger and higher-income adults showed limited adaptation. We also demonstrate that activity reallocation patterns differed across data-driven activity phenotypes (“Morning-,” “Neutral-,” and “Evening Walker”). These disparities suggest that structural factors (e.g., rigid work schedules, perceived safety) may constrain the capacity to flexibly adapt to time shifts for some populations. Physiologically, resting heart rate showed subtle intraday shifts mirroring behavioral changes, though differences were clinically insignificant. Our study provides the first large-scale causal analysis of DST’s influence using continuous wearables data, illustrating how observational data can generate real-world evidence to inform health-relevant policies.

## Introduction

Daylight Saving Time (DST) was first implemented in 1918 as a means to conserve energy by aligning waking hours with daylight. Over a century later, DST remains practiced in over 70 countries, yet it has become increasingly controversial. Recently, multiple countries have abandoned seasonal clock changes altogether, while repeated legislative efforts in the United States sought to make daylight saving time permanent^2^. This sustained political and public debate underscores the need for population-scale empirical evidence to inform policy decisions^3^.

A growing consensus, including position statements by the American Academy of Sleep Medicine, advocates for abolishing seasonal time changes in favor of permanent standard time^4^. The misalignment between biological rhythms and external time cues from DST has been linked to sleep disruption, circadian misalignment, and increased risks of myocardial infarction, ischemic stroke, and mood disorders^5–8^, with some reporting differential effects observed in spring vs fall transitions^2,5,6,8,9^. However, proponents argue that DST generates behavioral benefits, particularly by creating additional opportunities for outdoor physical activity by shifting evening light later. Based on common daily activity routines, the shift in time has been estimated to substantially increase opportunities for outdoor leisure among adults and children^10^. Because even modest increases in regular physical activity are known to confer measurable health benefits, DST-related behavioral changes could translate to meaningful implications for population health^11,12^. Resolving the debate around DST for public health benefits requires reliable, causal evidence that can evaluate both the behavioral and physiological responses to DST, yet such evidence remains limited.

Early studies on DST and health-related behaviors have relied on large-scale administrative data sources, such as hospital admission registries, health insurance claims, traffic crash databases, or self-reported surveys like the American Time Use Survey (ATUS)^7,13–16^. Using ATUS data from 2003–2009, Zick reported no detectable change in total time spent in moderate-to-vigorous physical activity among adults residing in Arizona, Colorado, New Mexico, and Utah following DST transitions^16^. Rosenberg and Wood, drawing on a telephone survey in Australia, found that nearly half the study participants reported perceived changes in time spent in physical activity during a typical week under daylight saving conditions^17^. While these studies have provided early insights into the impact of DST on physical activity patterns, their reliance on self-reported surveys and infrequent measurement limits their ability to precisely capture the day-to-day impact of DST on physical activity behavior and physiology. More recent work has begun to leverage wearable devices to move beyond self-reported measures and continuously quantify health-related behaviors and physiology in daily living settings. Goodman et al. used data from accelerometers (N=439) to demonstrate that longer daylight is associated with increased moderate-to-vigorous physical activity in children living in Europe and Australia, whereas no consistent changes were observed in the US pediatric population^18^. Heacock et al. found associations between DST transitions and significant shifts in sleep timing, consistency, and duration among wrist-worn fitness tracker users (N=24,000)^19^. Additional smaller-scale studies have used wearables to examine cardiovascular responses and light exposure changes during DST^20,21^. Despite these progresses, existing wearable-based studies have largely remained descriptive and depend on strict modeling assumptions (e.g., no unobserved confounders); because they rely on simple before-and-after comparisons without robust control groups, rigorous causal claims cannot be made.

In this study, we address these gaps by taking advantage of the natural variation over time and geography in one of the largest continuous wearable datasets available, the *All of Us* Research Program ^22,23^, spanning diverse demographic groups across the US. Arizona, the only state in the contiguous US that does not observe DST, serves as a natural control group, enabling a comparison to neighboring DST-observing Mountain Time states (Colorado, New Mexico, and Utah). Compared to previous studies that relied on simple before-and-after comparisons^16,18,19^, we employ a difference-in-differences (DiD) analysis that leverages this natural variation. Because DiD methods do not require adjusting for any individual-level confounders, our analysis relies on weaker assumptions that enable more rigorous causal inference. We then stratify our analysis by demographics and physical activity phenotypes to reveal heterogeneous effects across the population, providing the first large-scale causal analysis of DST’s influence on behavior and physiology from continuous wearable data.

## Results

### Cohort Description

The *All of Us* Research Program collected Fitbit data via two pathways: the Bring-Your-Own-Device (BYOD) program, in which participants shared retrospective Fitbit data, and the *All of Us* WEAR Program, which prospectively collected data beginning in March 2021 using study-provided devices. Our analytic cohort comprised 1,157 participants located in Arizona, Colorado, New Mexico, and Utah with valid intraday heart rate and step data for the two weeks surrounding each spring and fall DST transition during 2021-2023 (Supplementary Fig. 1)^24^. Of these, 66% (N=763) were enrolled through the BYOD program and 34% (N=394) through the *All of Us* WEAR program. Participants represented diverse demographic groups across the four states (Table 1).

**Table 1.** Demographic characteristics of participants in the final analytic cohort.

|  | Subgroup | Count (%) |
| --- | --- | --- |
| **Overall** | Total N | 1157 |
| **Age** | 18-44 | 268 (23.2%) |
|  | 45-64 | 406 (35.1%) |
|  | ≥ 65 | 483 (41.7%) |
| **Biological Sex** | Female | 790 (68.3%) |
|  | Male* | 350-375 (30.3-32.4%) |
|  | Unknown* | ≤ 20 (≤ 1.7%) |
| **Race** | Asian* | ≤ 20 (≤ 1.7%) |
|  | Black or African American | 26 (2.2%) |
|  | White | 986 (85.2%) |
|  | Other | 41 (3.5%) |
|  | Unknown* | 80-90 (6.9-7.8%) |
| **Ethnicity** | Hispanic or Latino | 123 (10.6%) |
|  | Not Hispanic or Latino | 1013 (87.6%) |
|  | Other/Unknown | 21 (1.8%) |
| **Annual Household Income** | <50k | 348 (30.1%) |
|  | 50-100k | 372 (32.2%) |
|  | >100k | 377 (32.6%) |
|  | Unknown | 60 (5.2%) |

*\*Counts below 20 are suppressed, and additional cells are obscured as needed to prevent back-calculation, complying with the All of Us Data and Statistics Dissemination Policy.*

### Behavioral response to DST: changes in physical activity

Visual inspection of hourly steps in Arizona (DST non-observing) and the surrounding Mountain Time states (DST-observing; Colorado, New Mexico, and Utah) revealed shifts in activity around the spring and fall transitions whereas Arizona showed no consistent change (Extended Data Fig. 1). After the “spring forward” transition, people in the DST-observing states were slower to start their activity in the mornings, particularly on Sunday (i.e., the morning of DST), and showed more activity in the late afternoon and evenings during the following days (Fig. 1A). In contrast, the “fall back” transition produced an opposite pattern: activity shifted earlier, with increased morning movement and reduced late-day steps in the DST-observing states (Fig. 1B).

**Figure 1.**
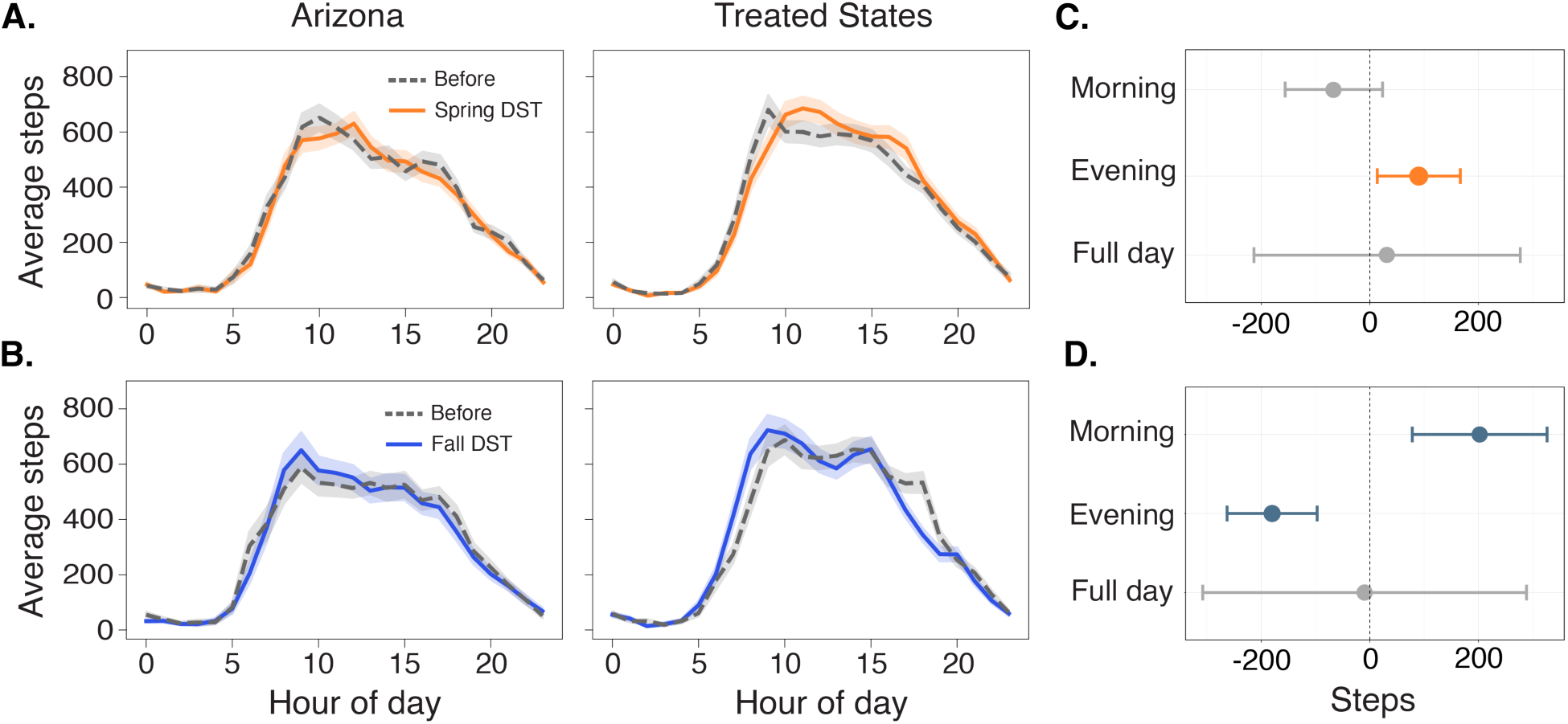
Behavioral effects of Daylight Saving Time (DST) transitions. **(A-B).** Average hourly step counts on the Sunday before and after the **(A)** Spring transitions in 2022 (N=812) and 2023 (N=1,072) and **(B)** Fall transitions in 2021 (N=820) and 2022 (N=837), combined. Shaded areas represent 95% confidence intervals. The near-parallel pre-transition trajectories across groups support the parallel trends assumption underlying the difference-in-differences (DiD) analysis. **(C–D)** Population-level DiD estimates for the effect of DST transitions on step counts across time-of-day periods (morning, evening, and full day) for **(C)** Fall transitions in 2021 and 2022 and **(D)** Spring transitions in 2022 and 2023. Points represent DiD coefficients, and error bars denote 95% confidence intervals. Effects that are not statistically significant (p ≥ 0.05) are displayed in grey.

Our DiD analysis corroborated these qualitative patterns and revealed contrasting effects between fall and spring DST transitions on intraday activity. Throughout, we report the results of a t-test on the estimated treatment coefficient from the DiD regression (see Methods: Difference-in-Difference), with standard errors clustered at the subject-level. DST transitions did not change total daily activity levels but rather restructured *when* people were active. Total daily steps did not alter significantly after either of the transitions (spring: β = 31.9, CI = [−214, 277], p = 0.799, t-test; fall: β = −9.5, CI = [−308, 289], p = 0.950, t-test). Yet, both transitions produced significant changes in activity distribution across the day, with opposing directional effects that mirror the shift in daylight availability. In spring, when sunset is delayed by an hour, evening activity slightly increases by 90 steps (CI = [14, 167], p = 0.021) (Fig. 1C) while morning activity showed a non-significant reduction (β = −66, CI = [−156, 24], p = 0.148). In fall, when sunrise is earlier, morning steps increased by 202 steps (CI = [78, 326], p = 0.001), roughly equating to about one and a half city blocks, while evening steps declined by 180 steps (CI = [−263, −97], p < 0.001) (Fig. 1D). This significant reciprocal change observed in the fall suggests that people respond to DST transitions by reallocating their activity timings (i.e., earlier movement) rather than by changing their overall activity level.

These intraday activity shifts were robust to the choice of post-transition observation window (see Methods: Sensitivity Analyses). Re-estimating treatment effects using post-transition periods ranging from one to six weeks produced qualitatively similar patterns (Supplementary Fig. 5), indicating that activity redistribution was not driven by a specific analysis window. The only attenuation observed was for fall morning activity, where the initial increase of 202 steps (CI = [78, 326], p = 0.001) was no longer statistically significant beyond the first post-transition week. Changes in evening activity following both spring and fall transitions persisted for up to six weeks.

We explored whether these temporal shifts varied by an individual’s typical activity timing. Based on steps data from six weeks preceding each transition, participants who consistently walk more in the morning than evening were labeled as “Morning Walkers;” participants who consistently walk more in the evening than in the morning were labeled as “Evening Walkers;” and participants who did not demonstrate differential levels of activity during any specific time period were labeled as “Neutral Walkers” (see Methods: Activity Phenotypes). We hypothesized that daylight shifts that align with a person’s preferred activity time would amplify activity levels. In particular, we expected this amplification for Evening Walkers in the spring since existing literature suggests evening hours represent a flexible, opportunistic window for leisure^18^.

The hypothesis held true for Morning Walkers in the fall (Fig. 2). When daylight moved earlier, they actively capitalized on the change, significantly increasing their morning activity (β = 636, CI = [146, 1126], p = 0.011) while reallocating some movement away from the evening (β = −144, CI = [−267,-21], p = 0.022). Interestingly, the reciprocal during the spring did not hold true for the Evening Walkers (i.e., they did not increase their evening steps), revealing an asymmetry in behavioral response in Morning vs Evening Walkers. We expected that the “spring forward” change, by providing more evening daylight, would create an opportunity for more physical activity in Evening Walkers. Despite the daylight now being better aligned with their typical active times, Evening Walkers did not show a significant increase in evening steps (β = 39.3, CI= [−166, 245], p = 0.708).

**Figure 2.**
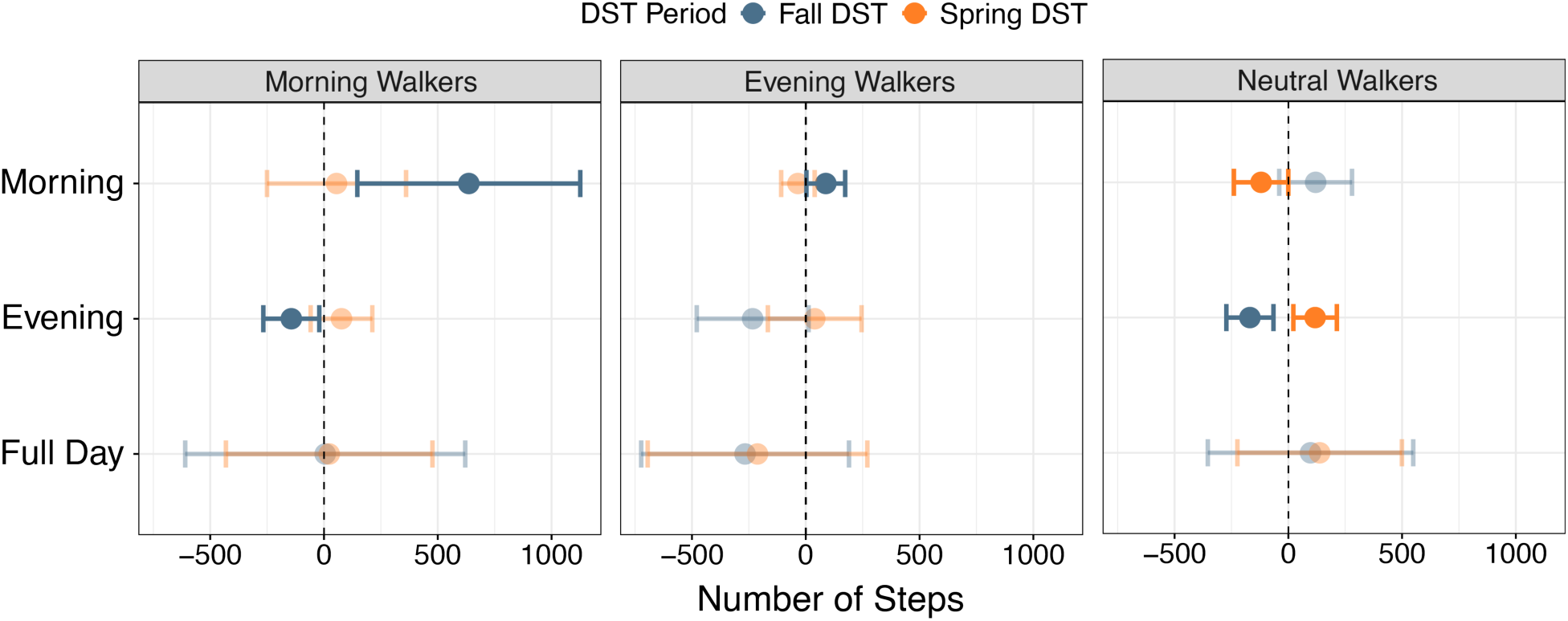
Heterogeneity in behavioral responses to Daylight Saving Time (DST) by physical activity phenotype. Effects stratified by activity phenotype (Morning, Evening, and Neutral Walkers) based on pre-DST activity timing classifications. Points represent difference-in-differences coefficients, and error bars denote 95% confidence intervals. Effects that were not statistically significant (p ≥ 0.05) are displayed with reduced opacity.

Neutral Walkers, who lack a strong temporal preference, primarily reallocated their activity across the day rather than amplifying activity in a specific time window. In fall, Neutral Walkers had a slight decrease in evening activity (β = −169, CI= [−272, −65], p = 0.001) with a non-significant increase in morning steps (β = 120, CI = [−40, 280], p = 0.142), behaving more like Morning Walkers. In the spring, neutral walkers took fewer steps in the darker morning (β = −119.8, CI = [−239, −0.2], p=0.05) and compensated with an almost identical increase in the lighter evening (β = 118, CI= [22, 213], p = 0.016), behaving more like Evening Walkers. Stratification by perceived neighborhood walkability revealed that this reciprocal morning-evening reallocation was evident among Neutral Walkers perceiving their neighborhood as more walkable (Extended Data Fig. 2). Across the four walkability features examined—perceived availability of transit, sidewalks, bicycle facilities, and recreational facilities—high transit availability was associated with the largest shift in activity following the spring transition, with decrease (β = −239, CI = [−402, −77], p = 0.003) and increase (β = 234, CI = [98, 370], p < 0.001) in the morning and evening steps, respectively (Extended Data Fig. 2A). Neutral Walkers reporting low perceived walkability showed no evidence of reciprocal morning-evening substitution following either DST transition (Extended Data Fig. 2B).

We also observed that the ability to reallocate intraday activity following the DST transition varied by age, household income, and sex (Extended Data Fig. 3). During the fall transition, adults aged 65 and older increased their morning steps (β = 281, CI = [80, 483], p = 0.006) while reducing their evening steps (β = 227, CI = [−341, −113], p < 0.001) (Extended Data Fig. 3A). In contrast, younger adults (18-45) exhibited no significant changes in either morning (β = 209, CI = [−58, 476], p = 0.124) or evening steps (β = −90, CI = [−286, 105], p = 0.365). This divergence may suggest that older adults are more flexible in adapting their daily schedules and more apt to follow daylight availability. Younger adults, by contrast, may be more constrained by fixed social schedules that limit temporal adjustments in their activity. This pattern is also directly reflective of the underlying distribution of activity timing preferences. The 65+ age group contains a substantially higher proportion of habitual Morning Walkers (36% vs. 8-18% in younger adults), which is also consistent with documented age-related shifts toward morning chronotypes^25^. As established in our primary analysis, individuals with pre-existing morning activity patterns capitalize on additional morning daylight, while those without such patterns show no to little increase in morning steps (Fig. 2).

Evening steps declined universally across income groups during fall (low-income: β = −207, CI = [−353.2, −61.4], p = 0.005; middle-income: β = −202, CI = [−369, −34], p = 0.018; high-income: β = −153, CI = [−287, −19], p = 0.025) (Extended Data Fig. 3B). However, only the lowest income bracket (≤ $50,000) compensated this decline in evening activity with increased morning steps (β = 335, CI = [100, 570], p = 0.005), whereas middle- and high-income groups showed no changes in morning activity (middle: β = 237, CI = [−2, 476], p = 0.052; high: β = 60, CI = [−128, 249], p = 0.532). The distribution of Morning vs Evening Walkers was similar across income groups, suggesting that income level does not play a role in activity phenotype. Instead, the morning increase among lower-income adults may reflect greater schedule flexibility— encompassing retirees or those with flexible work schedules (e.g., part-time, self-employed, gig, and/or mainly outdoor workers)—who continue their pre-DST behavior or reallocate activity toward daylight. In contrast, higher-income individuals may experience stronger occupational timing constraints, limiting their capacity to adjust morning behavior despite additional daylight or personal preference.

The spring transition, which shifts daylight toward evening hours, revealed an interesting difference by sex (Extended Data Fig. 3C). Females increased evening steps (β = 158, CI = [62, 253], p = 0.001), whereas males showed no significant change (β = −61, CI = [−185, 64], p = 0.339). Females may benefit from extended evening light induced by spring DST, reducing pre-existing barriers to evening activity (e.g., perceived outdoor safety)^26^.

### Physiological response to DST: changes in average resting heart rate

Across both DST transitions, changes in average resting heart rate (RHR) were small (ranging from −2 to +2 beats per minute (bpm)), differences which are unlikely to be clinically meaningful (Extended Data Fig. 4). In fall, RHR during the evening declined by 0.8 bpm (β = −0.8, CI = [−1.4, −0.2], p = 0.014) while morning RHR rose slightly (β = 1.3, CI = [0.7, 2.0], p < 0.001). In spring, RHR declined in the morning (β = −1.0, CI = [−1.6, −0.5], p < 0.001) without any changes in the evening RHR (β = 0.4, CI = [−0.1, 1.0], p = 0.127). We found no significant differences in daily average RHR during either seasonal transition (spring: β = 0.0, CI = [−0.3, 0.4], p = 0.763; fall: β = −0.1, CI = [−0.5, 0.3], p = 0.621).

## Discussion

We used intraday Fitbit data from over 1,000 participants in the *All of Us* Research Program to investigate the behavioral and physiological effects of DST policy. Our DiD analysis reveals that while DST transitions influence *when* people are active, they do not significantly alter how much they are active. Our primary finding refutes the common argument that increased evening light due to DST creates more activity in the US^10^. In line with previous work, we found no significant net gain or loss in total daily steps following either the spring or fall transitions^10,12^. The one-hour shift in daylight instead triggered a significant temporal reallocation of physical activity (Fig. 1). This was most evident in the “fall back” transition, which prompted a clear reciprocal shift, with a 202-step increase in the morning and a 180-step decrease in the evening (Fig. 1D).

A common intuition in DST-related physical activity research is that extended evening daylight creates additional opportunities for discretionary movement^10^. Our findings suggest that while these opportunities do emerge, they do not translate into a net increase in physical activity. Although evening steps increased following the spring transition (Fig. 1), total daily step counts remained unchanged, even among Evening Walkers whose typical activity patterns were most closely aligned with the newly illuminated hours (Fig. 2). These results challenge the assumption that evening light functions as an effective intervention for increasing total daily activity levels. Rather, while light availability may influence the timing of movement, it does not inherently generate the motivation or capacity to expand the overall volume of activity.

Prior investigations into DST have documented increases in pedestrian and cycling activity following transitions that extend daylight, suggesting that ambient light promotes active travel^27,28^. Separately, a large body of work has shown that walkable built environments support higher levels of routine walking, particularly for transport and commuting^29–31^. These findings motivate the expectation that DST-related increases in pedestrian activity might be most evident in highly walkable settings, where opportunities for active travel are greatest. Our results, however, do not support this hypothesis. Neutral Walkers—those who do not exhibit preferred activity timing—did not increase their overall steps taken, even in areas perceived to be highly walkable (Extended Data Fig. 2A). Instead, they consistently reallocated existing activity to periods of greater ambient light. The reallocating pattern, however, was not observed in Neutral Walkers residing in lower perceived walkability areas. This suggests that DST-related increases in pedestrian activity reported in prior studies may reflect temporal and spatial redistribution of routine movement—enabled by pedestrian infrastructure—rather than increases in overall physical activity.

Demographic stratification further reinforced that no subgroup achieves a net gain in total steps (Extended Data Fig. 3), but the pattern of reallocation varied by age, annual household income, and sex, suggesting distinct behavioral (e.g., rigid work hours) and structural considerations (e.g., perceived safety). Older adults, females, and lower-income individuals exhibited significant increases in morning steps and a decrease in evening steps following the fall transition. On the other hand, following the spring, females and ages 45-65 showed greater evening activity, whereas males primarily reduced morning movement. Yet, in all cases, these gains were offset by compensatory reductions at other times of day, resulting in a net-zero change in total activity volume.

Emerging evidence, particularly randomized crossover studies in which individuals switch the timing of physical activity while total volume is held constant, suggests that activity timing carries health relevance beyond total volume. These studies report time-of-day–specific physiological adaptations, including differential engagement of carbohydrate and lipid metabolic pathways^32^ and improvements in 24-hour glucose levels^33^. With larger randomized trials in more diverse populations now underway to rigorously evaluate the causal effects of exercise timing on cardiometabolic, sleep, and circadian outcomes, activity timing is increasingly recognized as a modifiable dimension of health behavior rather than a secondary characteristic^34,35^. Findings from these future crossover studies would help contextualize our results and evaluate the public health tradeoffs of DST policies.

Just as with physical activity, we found no net change in daily average RHR (Extended Data Fig. 4). However, we observed a similar pattern of intraday changes as physical activity. In fall, for instance, RHR rose slightly in the morning while declining in the evening. Although these acute changes were too small to be clinically meaningful, they demonstrate that the body’s rhythms are being subtly perturbed alongside behavioral shifts. In this analysis, we observed the immediate effects of DST transitions using a two-week observation window. Future studies can collect higher-resolution data to detect the more subtle physiological changes (e.g., heart rate variability) indicative of the circadian misalignments.

From a methodological standpoint, our approach utilizing large-scale wearable data establishes stronger causal evidence than prior observational research. While previous studies have examined DST effects, they have largely relied on either cross-sectional comparisons (which fail to account for baseline population differences) or simple pre-post analyses (which left open the possibility that observed changes reflected seasonal trends rather than DST itself)^16,18^. We employed a DiD approach that addresses these limitations and isolated the causal effect from concurrent seasonal confounders. In this framework, causal identification does not require explicit modeling of time-varying covariates as long as Arizona and its DST-observing neighbors would have experienced similar activity trajectories in the absence of DST transitions (i.e., parallel-trends assumption). We reduced the likelihood of violating this assumption by focusing on a short pre-treatment time window and neighboring Mountain Time states that share similar geography, latitude, and time zone, and thus experience nearly identical daylight patterns (Fig. 1). Weather conditions in this region also evolve in close synchrony—spring warming, fall cooling, and precipitation trends are highly aligned across Arizona, Colorado, Utah, and New Mexico. Because the DiD design is robust to individual-level confounding, our specification accounts for time-invariant climatic and behavioral differences. Moreover, given the shared seasonal conditions across these states, our model also absorbs region-wide weather shocks. Consequently, only idiosyncratic, state-specific anomalies coinciding precisely with the DST transition could bias our estimates, and–to our knowledge–no such shocks occurred during the study period.

Several limitations should be considered when interpreting these findings. Our sample skews toward female and higher socioeconomic demographics who likely have greater flexibility for physical activity (Table 1), suggesting our estimates may not represent the general US population^36–38^. In addition, sleep outcomes were not evaluated due to Fitbit’s unreliable sleep tracking^39^, especially during DST transitions, as reported by many Fitbit users. Given these data quality concerns and the absence of validated correction algorithms, we excluded sleep analyses to ensure methodological rigor, though future work could incorporate algorithmic calibration to enable more reliable sleep assessment. Our estimation of the behavioral effects through predefined time windows (e.g., morning, evening) may not fully capture the temporal shifts in physical activity; alternative approaches using hourly bins or rolling activity windows could provide finer resolution.

Our study leveraged large-scale wearable data from the *All of Us* Research Program to evaluate the behavioral effects of DST transitions in the United States. Contrary to the common assumption that DST would promote physical activity through added evening light, we found no evidence that DST amplifies total physical activity. Instead, DST primarily reallocated when people were active, producing reciprocal shifts between morning and evening hours. We also found that the ability to adapt to this shift is not uniform across the population. As policymakers continue to debate the future of DST, such findings should be interpreted cautiously and weighed against the well-documented disruptions DST causes to sleep, circadian rhythms, cardiovascular health, and public safety^2,40–47^.

## Methods

### Data from the *All of Us* Research Program

We used the demographic, survey, and Fitbit data from the NIH’s *All of Us* Research Program^16^. Our analysis used the most up-to-date Controlled Tier dataset version 8 (C2024Q3R5), which includes non-date-shifted intraday Fitbit data (through October 1, 2023) and the first three digits of participant ZIP codes.

From a total of 59,018 participants who shared any Fitbit data, we identified 53,812 individuals who contributed intraday heart rate data (Supplementary Fig. 1). Of these, 51,364 participants also had both state and ZIP codes. After excluding individuals with discrepancies in their geolocation information, we retained 4,375 participants across Arizona, Colorado, New Mexico, or Utah. To focus on the immediate effects of DST, we included participants with valid data on any day during the first week of spring or fall DST transition for years 2021, 2022, and 2023. After applying our inclusion criteria, our cohort included 1,531 unique individuals with both intraday HR and steps data (Supplementary Fig. 1). Year-by-year transition breakdown is shown in Extended Data Table 1.

### Data Processing

To address missingness from inconsistent device wear, we defined “valid days” using sensitivity thresholds ranging from 0% to 100% daily wear or wear during specific time windows (e.g., morning and evening) (Supplementary Fig. 3A). Wear time was calculated as the sum of all minute-level HR data available for each person per day; wear threshold was defined as the proportion of observed wear time over the potentially observable data^24^. For example, the potentially observable data for a full day is 1440 minutes (60 min/hour * 24 hour/day). We required participants’ average wear time for each DST period to meet wear-time thresholds (75%) to enable within-person temporal comparisons of the specific time window. We additionally performed sensitivity analysis with different wear threshold choices (ranging from 75% to 95%), illustrating the robustness of our analysis to this hyperparameter (Supplementary Fig. 3B & 3C).

To reduce the influence of extreme activity values, we excluded person-days with step counts exceeding the 99.9th percentile within each time window (morning, evening, night, or full day). Fewer than 20 participants were removed following this criterion. From 1,531 unique participants, our final analytic cohort included 1,157 participants (Supplementary Fig. 1). We summarize the demographics of the full analytic cohort residing in the four states and who had valid data in any of the four DST transitions (Table 1). Step counts stratified by Arizona and Mountain Time states are shown in Supplementary Fig. 4. Of note, some individuals contributed data to multiple transitions, while others participated in only one.

Significant effects of DST in those with flexible walking patterns (i.e., Neutral Walkers) were observed (Fig. 2). We therefore examined whether built environment characteristics moderated the behavioral impact of DST within this group. We selected five questions from the *All of Us* Research Program’s Social Determinants of Health survey to assess perceived neighborhood walkability (Supplementary Table 1). The survey was distributed to all enrolled participants as a one-time voluntary questionnaire between November 2021 and September 2023. Questions assessed perceptions of amenities and infrastructure within participants’ built environment, defined as a 10-15 minute walk from their residence. Items covered environmental infrastructure (e.g., presence of sidewalks, bicycle facilities, transit stops) and accessibility (e.g., availability of recreational facilities). Response options that included “Don’t know” or “Does not apply” were treated as neutral midpoint values (e.g., neither agree nor disagree) to preserve ordinal structure while minimizing data loss, while questions skipped were considered missing. We then binarized responses as high (score > 3) vs low accessibility (score ≤ 3) for each item.

### Feature Extraction

Behavioral features were derived from step data, calculated as total steps per day and across specific time windows. We chose resting heart rate (RHR) as the physiological feature of interest to minimize confounding influences from physical activity. RHR was calculated as the average heart rate during restful periods (steps = 0). We examined average RHR across the full day and within time-specific windows.

### Activity Phenotypes

We estimated the effect of DST on individuals with different activity patterns by defining activity phenotypes based on step count data collected during the six weeks preceding the transition (Supplementary Fig. 5; Supplementary Table 2). Participants were classified as Morning, Evening, or Neutral Walkers using a rule-based approach. For each day within this period, we compared the number of steps taken in the morning versus the evening. Days with more morning steps were assigned a score of +1, days with more evening steps were assigned −1, and days with no clear preference were assigned 0. Each participant’s mean daily score over the final six days before DST represented their time-of-day tendency. Participants with average scores > 0.5 were classified as Morning Walkers, those with scores < –0.5 as Evening Walkers, and those between –0.5 and 0.5 as neutral walkers. This classification required participants to have at least one day of both morning and evening step counts with ≥75% wear level. The distribution of Morning, Evening, and Neutral Walkers is illustrated in Supplementary Fig. 5.

### Difference-in-Difference (DiD)

Our analysis leverages state-level differences in DST observance. Specifically, because Arizona (with the exception of the Navajo Nation) does not participate in the time change, its residents are not exposed to DST changes^16^. We leveraged this natural variation over time and across states to conduct a difference-in-difference (DiD) analysis using Arizona residents as our “control” group.

The DiD analysis is robust to time-invariant omitted variables that are not accounted for in other DST studies, such as Goodman et al.’s accelerometer-based observational analyses in children that attribute higher MVPA to longer evening daylight in Europe and Australia without a geographic counterfactual control, and Ferguson et al.’s within-person pre/post wearable study in adults that detects only small shifts (e.g., +8 min/day sedentary after DST ends) but lacks a no-DST control to separate seasonal effects^18,48^. Furthermore, while Zick employed a similar geographic comparison using Arizona, pre-DST baseline data were absent in their approach^16^.

We assessed the pre-treatment trends by plotting the average hourly step counts during the week before and after the DST transition, separated by day of the week and control vs intervention (Fig. 1A & 1B). The pre-DST patterns (grey) for Arizona and the Mountain Time states follow highly similar shapes and magnitudes, indicating parallel pre-transition trends in activity.

Our DiD analysis controls for day-of-week and yearly behavioral differences^49^. The regression function is:

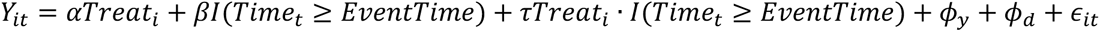

where *Y_it_* measures the outcome for person *i* at time *t*, *Treat_t_* is a binary indicator for whether person *i* is in a region of DST exposure, I(*Time_t_* ≥ *EventTime*) indicates whether the corresponding time period is before or after the time change, *ϕ_y_* is a matrix of year-level fixed effects, *ϕ_d_* is a matrix of day-of-week fixed effects, and *ɛ_it_* is random and clustered by subject. Throughout the paper, we report the estimated treatment coefficient from the DiD regression along with the p-value from the t-test.

We estimate the effect of DST on the mean RHR or total step counts in three time-windows: (1) morning (6-9AM), (2) evening (5-7PM), and (3) full day (24 hours) (Fig. 1 and Extended Data Fig. 4). We fit separate DiD regressions for fall and spring DST changes and time window (morning and evening). Analyses are conducted using the statsmodels package in Python.

### Sensitivity Analyses

To test the robustness of our findings to key analytic choices, we conducted a series of sensitivity analyses. First, we repeated the primary analyses under alternative wear-time thresholds, ranging from 75% to 95%, demonstrating that the estimated effects are stable across this hyperparameter choice (Supplementary Fig. 3). Second, while our main results focus on the effects estimated over the first week following the time change to capture short-term impacts, we assessed the persistence of DST effects over longer horizons to evaluate the robustness to the choice of our analysis window. Specifically, we re-estimated the treatment effect using the same DiD model specification but with longer post-transition windows, ranging from one to six weeks, yielding qualitatively similar results (Supplementary Fig. 2).

### Heterogeneous effects of DST

We examined heterogeneity in DST effects by activity phenotype (Fig. 2), perceived walkability measures (Extended Data Fig. 2), and by demographics, including income level, biological sex, and age group (Extended Data Fig. 3). We estimated separate DiD regressions for each combination of subgroup and time period. Only individuals with non-missing information for that subgroup were included in these analyses.

## Data availability

To ensure participant privacy, data used for this study is available to approved researchers following registration, completion of ethics training, and attestation of a data use agreement through the *All of Us* Research Workbench platform, which can be accessed via https://workbench.researchallofus.org/login.

## Acknowledgements

This project is supported by NSF CAREER Award (#2339669). H.J. is supported by the National Institutes of Health (NIH) National Heart, Lung, and Blood Institute F31 Fellowship (F31HL179990-0); S.K is supported by the Apple Scholar in AI/ML Fellowship. We gratefully acknowledge *All of Us* participants for their contributions, without whom this research would not have been possible. We thank the NIH’s *All of Us* Research Program for making available the participant data and the cohort examined in this study. This work was reviewed and submitted to the Machine Learning for Health (ML4H) 2025 Findings Track (non-archival), and we gratefully acknowledge the constructive feedback provided by four anonymous reviewers. We also thank Sarah Jiang for her contributions to the initial literature review during the early stages of this work.

## Author contributions

H.J. and K.W. conceived the initial idea for the study. H.J., S.K., and K.W. developed the research framework, and J.D. supervised the project. K.W. performed the literature review, and H.J. curated the data for analysis. S.K. conducted the formal analysis under the supervision of A.V. The results were interpreted collaboratively by all authors. H.J., S.K., and K.W. drafted the initial manuscript. All authors contributed to data interpretation, critically revised the manuscript, and approved the final version for submission.

## Competing Interests

J.D. sits on the Google Consumer Health Advisory Board and is a consultant to Samsung Research America. All other authors declare no competing interests.

## Materials & Correspondence

Correspondence to Jessilyn Dunn

## Notes

### Author Declarations

Data used for this study is available to approved researchers following registration, completion of ethics training, and attestation of a data use agreement through the *All of Us* Research Workbench platform, which can be accessed via https://workbench.researchallofus.org/login.

### Summary of Updates

Expanded literature review to more clearly situate our work within the broader field of Daylight Saving Time (DST) research and its relevance to health outcomes; additional sensitivity analyses incorporating extended post-transition windows into and out of DST; a new subgroup analysis examining how built environment characteristics, such as perceived transportation accessibility, may moderate the effects of daylight saving time on behavioral patterns

