## Extended Data for "Should We Keep Changing the Clock? Characterizing Causal Effects of Daylight Saving Time on Behavior and Physiology"

**Extended Data Table 1.** Availability of heart rate and step count data across Daylight Saving Time (DST) transitions for Arizona (does not observe DST) and Mountain Time (MT) states (observe DST).

| **Year** | **Region** | **Data availability** | **After Data cleaning** |
|---|---|---|---|
| **2023 Spring** | Total | 1072 | 787 |
| | Arizona | 424 | 312 |
| | MT States | 648 | 475 |
| **2022 Fall** | Total | 837 | 549 |
| | Arizona | 376 | 218 |
| | MT States | 461 | 331 |
| **2022 Spring** | Total | 812 | 594 |
| | Arizona | 417 | 290 |
| | MT States | 395 | 304 |
| **2021 Fall** | Total | 820 | 453 |
| | Arizona | 418 | 194 |
| | MT States | 402 | 259 |

**Extended Data Fig. 1. Average hourly step counts during the week before and after Daylight Saving Time (DST). A.** Average steps taken during spring transitions in 2022 and 2023. **B.** Average steps taken during fall transitions in 2021 and 2022. Treated States include Colorado, New Mexico, and Utah. Shaded areas represent 95% confidence intervals.

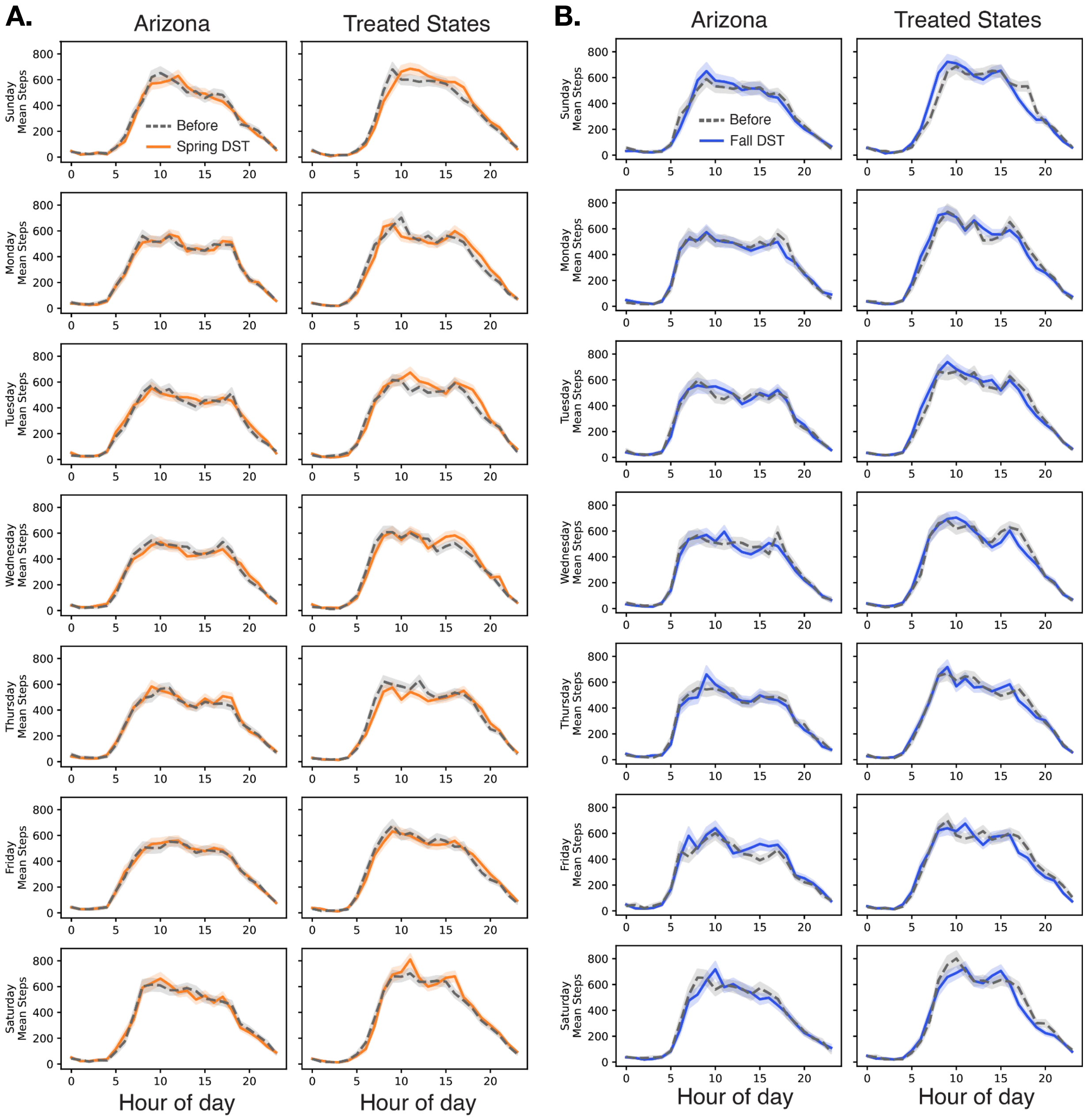

**Extended Data Fig. 2. Heterogeneous effects of DST transitions on step counts across walkability measures.** Difference-in-differences (DiD) point estimates with 95% confidence intervals for the effects of DST transitions on step counts across different time-of-day periods for Neutral Walkers, stratified by **(A)** high and **(B)** low availability of walkability-related features, including sidewalks, public transit, bicycle facilities, and recreational facilities. Estimates are shown for full-day, morning, and evening periods. Points denote estimated effects, horizontal bars indicate 95% confidence intervals, and the vertical dashed line denotes no change. Estimates that are not statistically significant ($p \geq 0.05$) are displayed with reduced opacity.

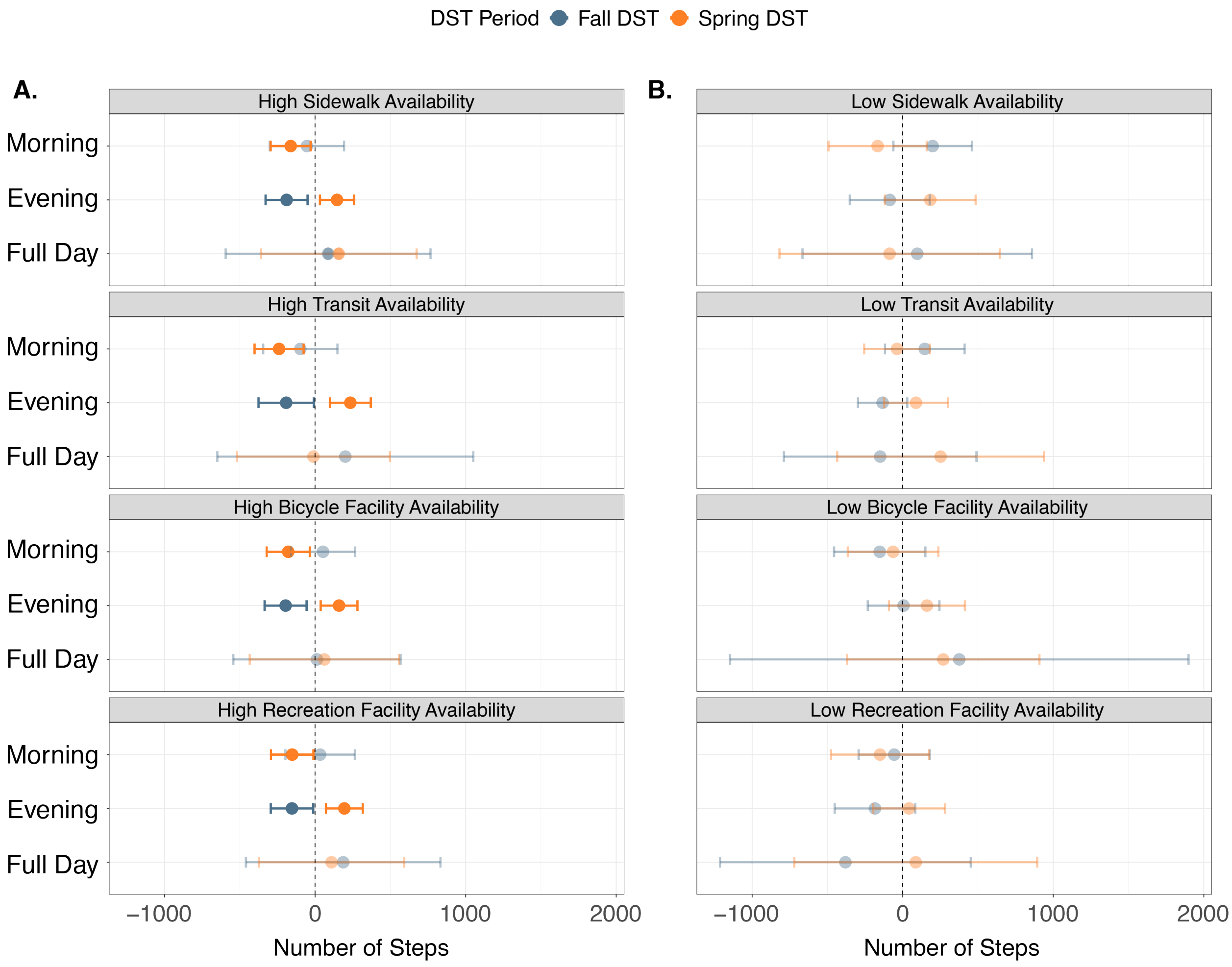

**Extended Data Fig. 3. Heterogeneous effects of Daylight Saving Time (DST) on step counts across demographic subgroups.** Difference-in-differences (DiD) point estimates with 95% confidence intervals across different time-of-day periods, stratified by demographic characteristics: **(A)** age group, **(B)** annual household income **(C)** sex. Effects that are not statistically significant (p ≥ 0.05) are displayed with reduced opacity.

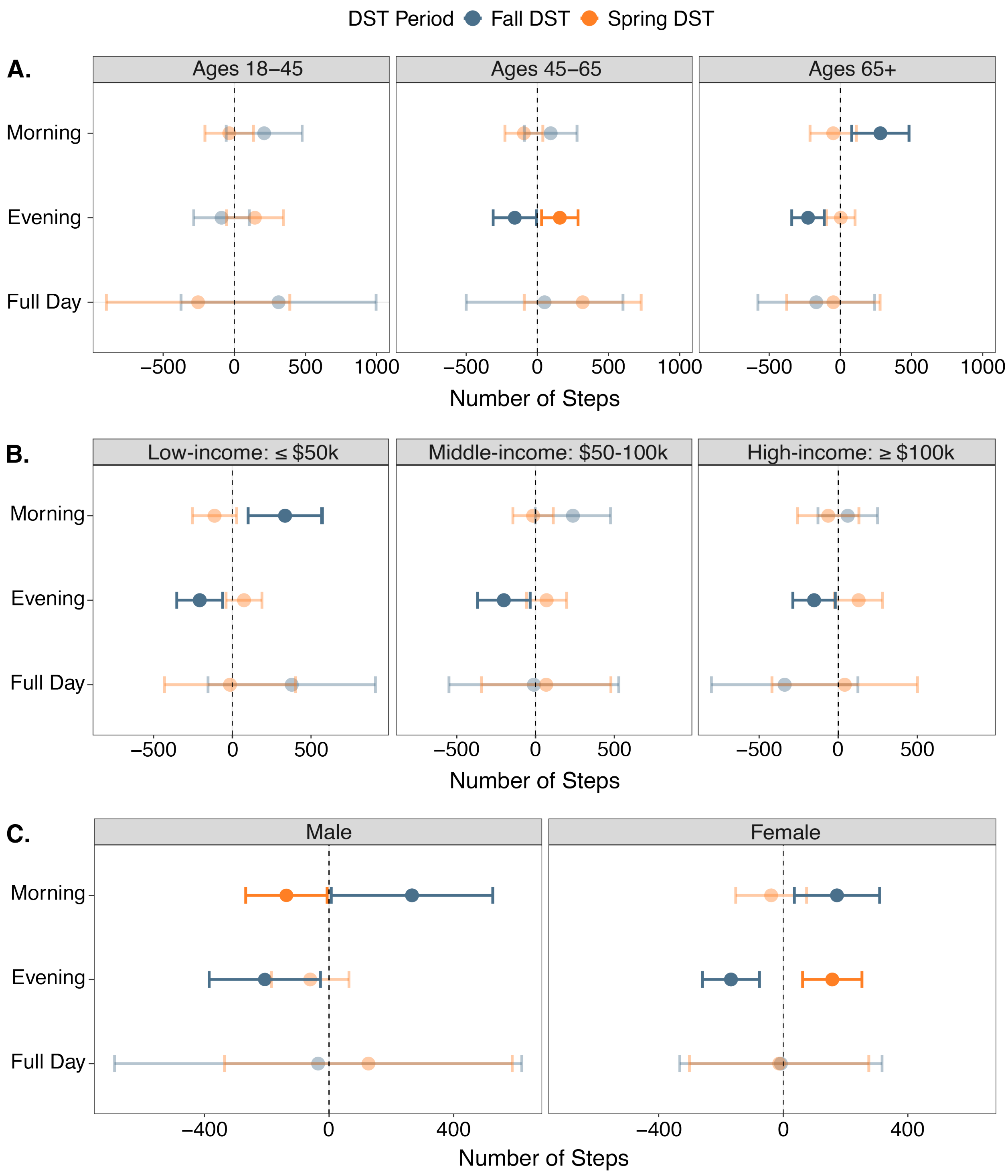

**Extended Data Fig. 4. Effects of Daylight Saving Time (DST) transitions on resting heart rate.** For each time-window, population-level difference-in-difference point estimates are reported with 95% confidence intervals. Effects that are not statistically significant ($p \geq 0.05$) are displayed with reduced opacity.

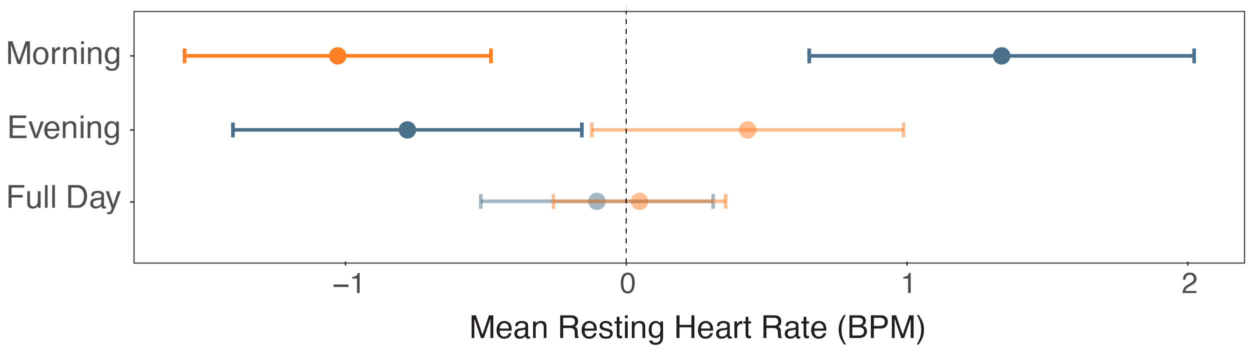
