## Supplementary Table for "Should We Keep Changing the Clock? Characterizing Causal Effects of Daylight Saving Time on Behavior and Physiology"

**Supplementary Table 1.** Completion rates and counts (N, %) of high (score > 3) and low walkability (score ≤ 3) among Neutral Walkers across Fall 2021 to Spring 2023.

| Question Prompt | | Spring 2023 | Fall 2022 | Spring 2022 | Fall 2021 |
|---|---|---|---|---|---|
| There are sidewalks on most of the streets in my neighborhood. | Completion rate | 354 (81.9%) | 238 (82.1%) | 257 (82.4%) | 204 (81.3%) |
| | High | 279 (78.8%) | 158 (77.7%) | 208 (80.9%) | 163 (79.9%) |
| | Low | 75 (21.2%) | 53 (22.3%) | 49 (19.1%) | 41 (20.1%) |
| It is within a 10-15 minute walk to a transit stop (such as bus, train, trolley, or tram) from my home. | Completion rate | 354 (81.9%) | 240 (82.8%) | 255 (81.7%) | 201 (80.1%) |
| | High | 239 (67.5%) | 151 (62.9%) | 166 (65.1%) | 132 (65.7%) |
| | Low | 115 (32.5%) | 89 (37.1%) | 89 (34.9%) | 69 (34.3%) |
| There are facilities to bicycle in or near my neighborhood, such as special lanes, separate paths or trails, or shared use paths for cycles and pedestrians. | Completion rate | 350 (81.0%) | 237 (81.7%) | 256 (82.1%) | 202 (80.5%) |
| | High | 290 (82.9%) | 194 (81.9%) | 209 (81.6%) | 174 (86.1%) |
| | Low | 60 (17.1%) | 43 (18.1%) | 47 (18.4%) | 28 (13.9%) |
| My neighborhood has several free or low-cost recreation facilities, such as parks, walking trails, bike paths, recreation centers, playgrounds, public swimming pools, etc. | Completion rate | 340 (78.7%) | 232 (80.0%) | 245 (78.5%) | 200 (79.7%) |
| | High | 279 (82.1%) | 189 (81.5%) | 190 (77.6%) | 163 (81.5%) |
| | Low | 61 (17.9%) | 43 (18.5%) | 55 (22.4%) | 37 (18.5%) |

**Supplementary Table 2.** Number of participants in each activity phenotype at each Daylight Saving Time (DST) transitions.

| | N (%) | | | |
|---|---|---|---|---|
| | **Spring 2023** | **Fall 2022** | **Spring 2022** | **Fall 2021** |
| **Neutral Walkers*** | 420-440 (53-56%) | 280-300 (51-55%) | 300-320 (51-54%) | 240-260 (53-57%) |
| **Morning Walkers** | 179 (22.7%) | 133 (24.2%) | 134 (22.6%) | 109 (24.1%) |
| **Evening Walkers** | 174 (22.1%) | 125 (22.8%) | 144 (24.2%) | 91 (20.1%) |
| **Unclassified*** | ≤ 20 (0-1%) | ≤ 20 (0-4%) | ≤ 20 (0-4%) | ≤ 20 (0-4%) |

**Note: Counts below 20 are suppressed, and additional cells are obscured as needed to prevent back-calculation, complying with the All of Us Data and Statistics Dissemination Policy.*
