## Supplementary Fig for "Should We Keep Changing the Clock? Characterizing Causal Effects of Daylight Saving Time on Behavior and Physiology"

**Supplementary Fig. 1. Flowchart of the cohort inclusion criteria**

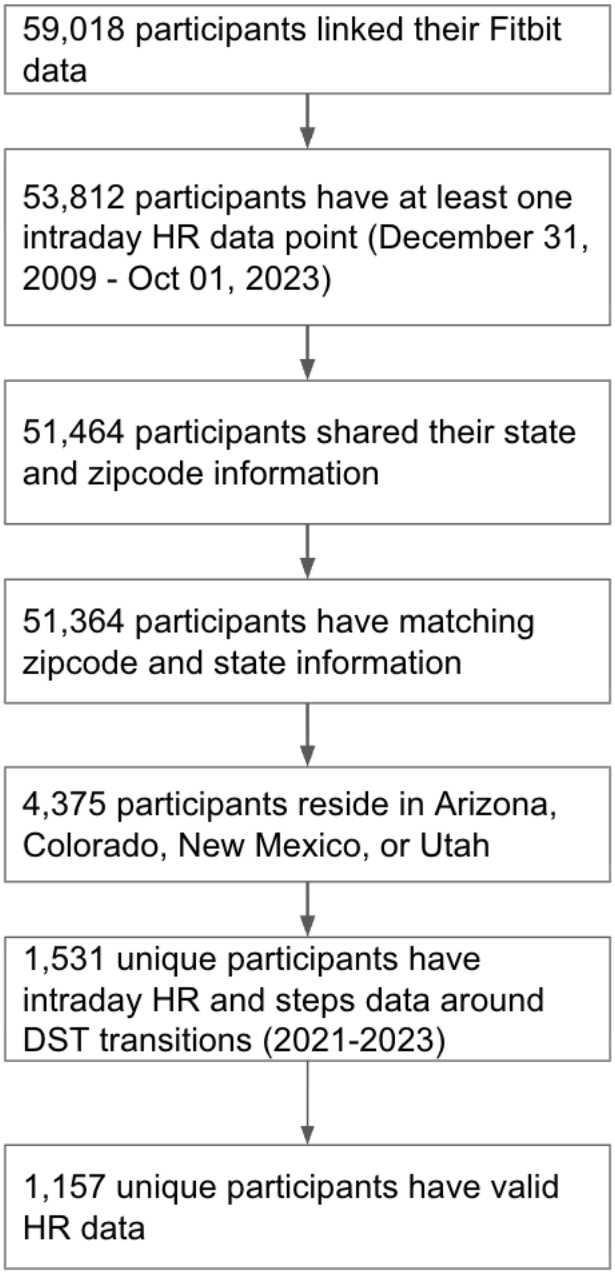

**Supplementary Fig. 2. Sensitivity of intraday activity shifts to the length of the post-DST observation window.** Estimated changes in step counts associated with daylight saving time (DST) transitions are shown for full-day, morning, and evening periods using post-transition observation windows ranging from 1 to 6 weeks. Points represent difference-in-difference (DiD) coefficients, and error bars denote 95% confidence intervals. Effects that are not statistically significant ($p \geq 0.05$) are displayed with reduced opacity. Across window lengths, the initial increase in morning activity following the fall transition attenuated with longer follow-up and lost statistical significance beyond shorter windows, whereas changes in evening activity following both fall and spring transitions remained persistent across extended observation periods.

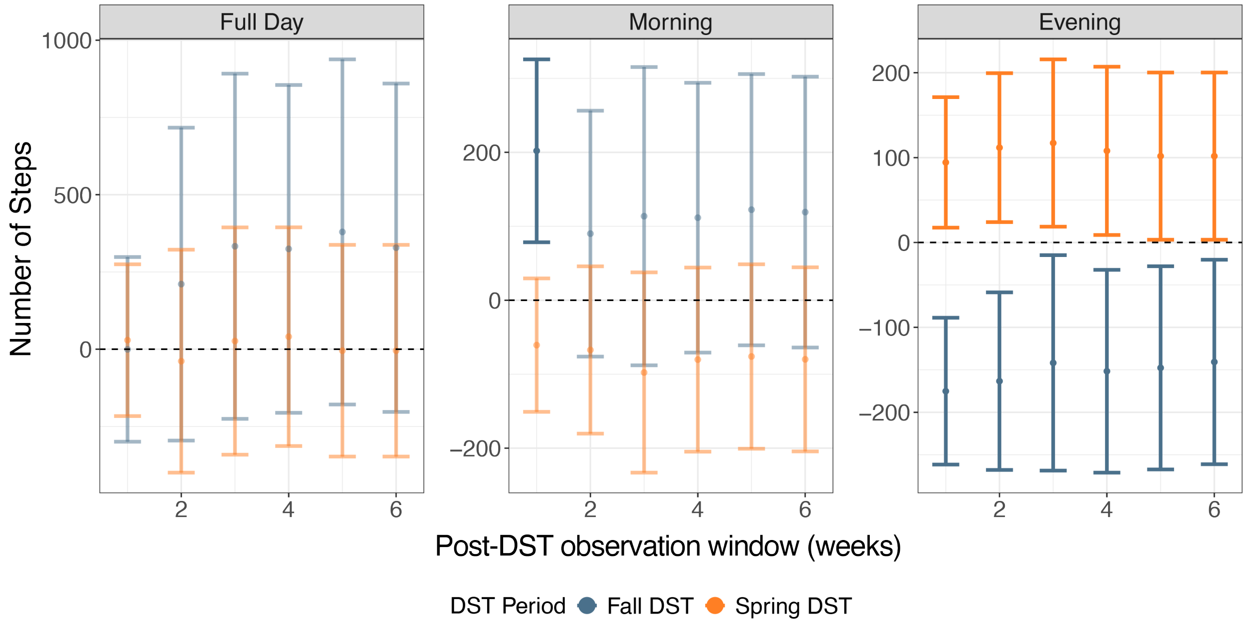

**Supplementary Fig. 3  Impact of missing data on the robustness of Daylight Saving Time (DST) effect estimates. (A)** Example of data missingness in Spring 2023 under varying wear-time thresholds, ranging from 0% to 100% within specific time windows. **(B-C)** Estimated difference-in-differences (DiD) point estimates with 95% confidence intervals (grey) on **(B)** Step counts and **(C)** Average resting heart rate when varying the minimum wear-time threshold required for inclusion. Results are largely consistent across wear thresholds, indicating robustness of the findings to differences in data missingness. Full-day estimates at the 0.95 threshold were excluded due to instability in the variance estimator.

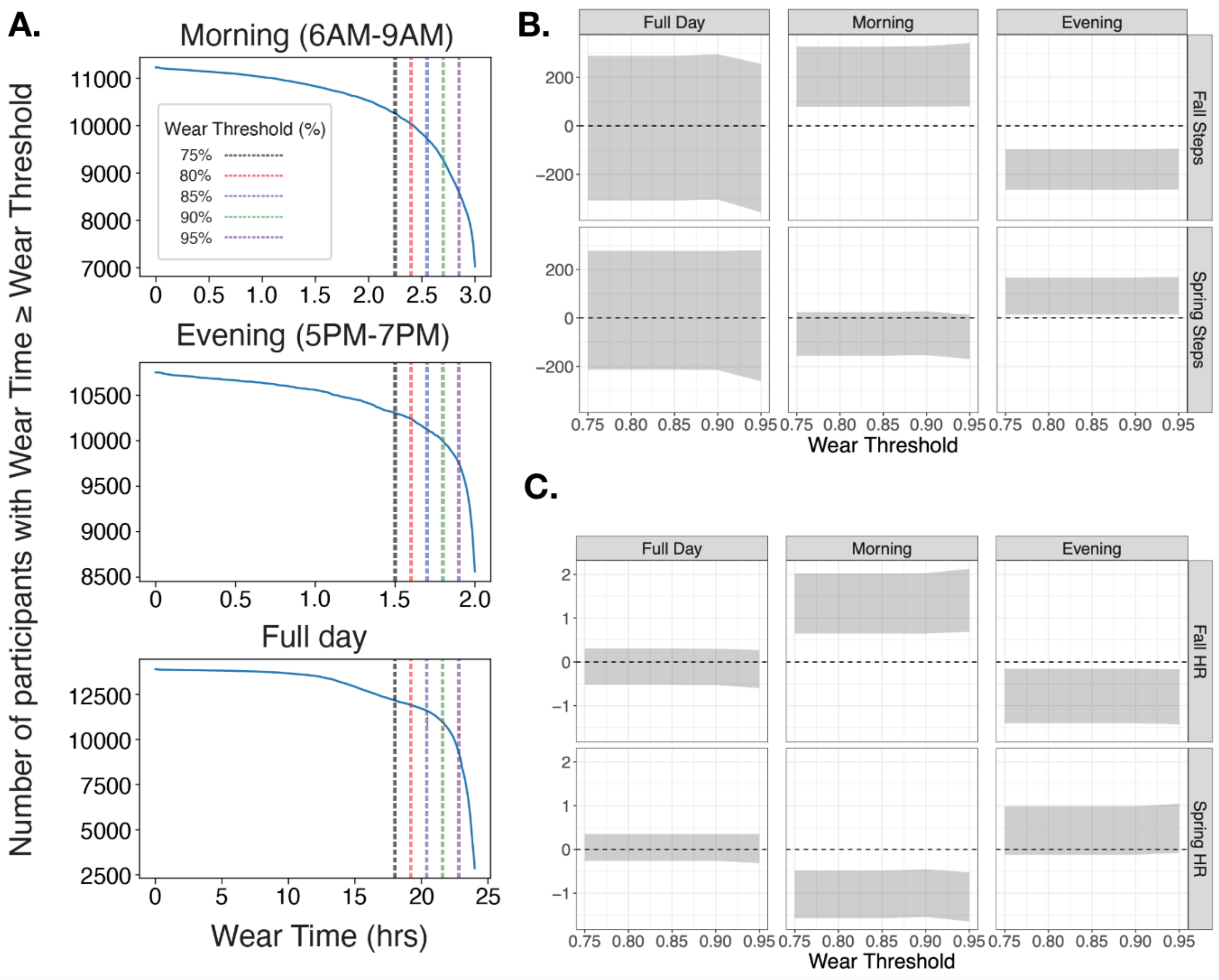

**Supplementary Fig. 4. Distribution of average daily step counts prior to daylight saving time (DST) transitions.** Boxplots show the distribution of mean daily steps per person, calculated over the six weeks preceding each DST transition, stratified by time of day (morning, evening, and full day), year, and region (Arizona vs. states observing DST). Points represent individual-level observations; boxes indicate the interquartile range with medians shown as horizontal lines, and whiskers denote 1.5× the interquartile range.

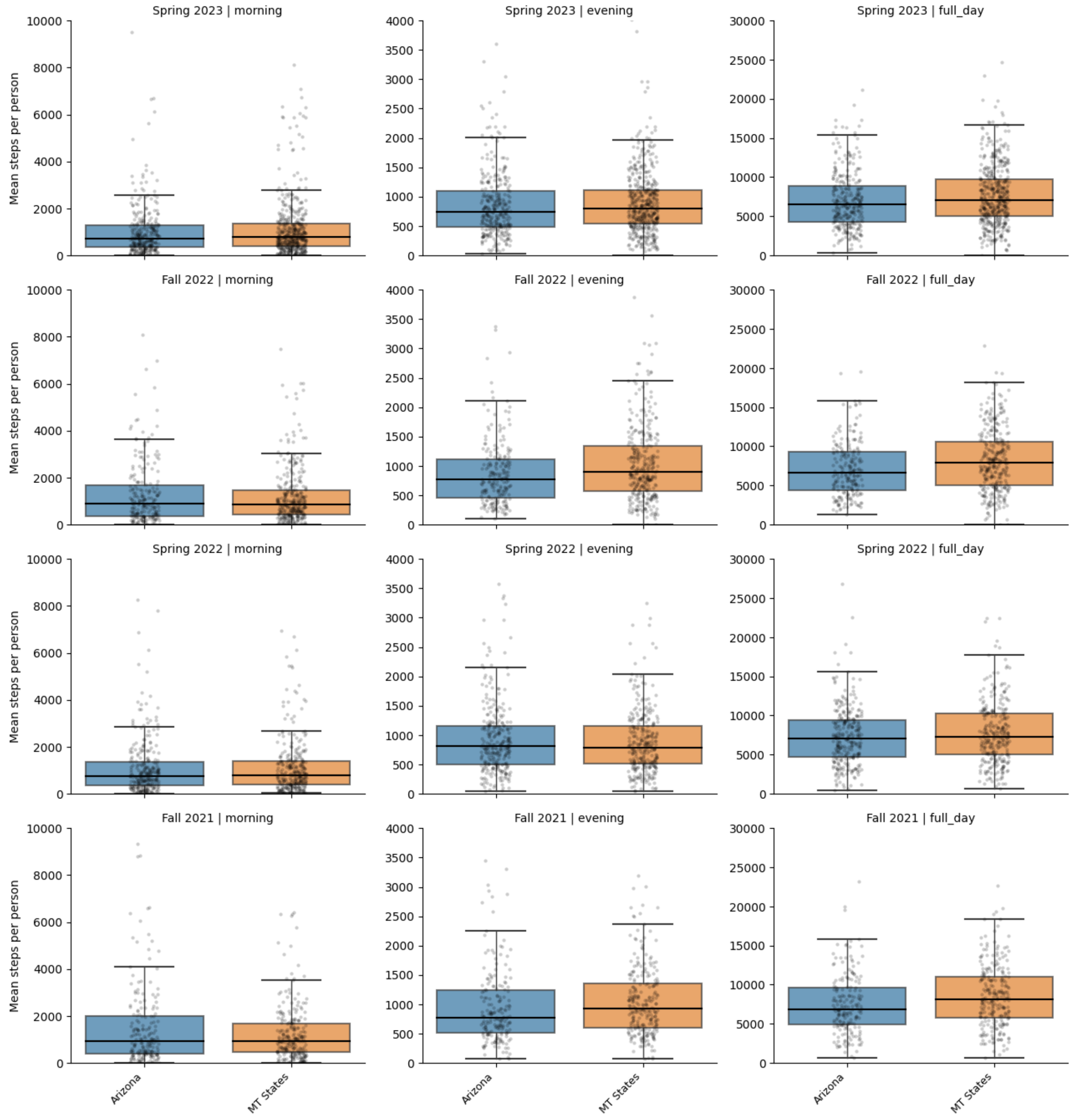

**Supplementary Fig. 5. Example of activity phenotypes using Spring 2023 data.** Over half of participants were classified as Neutral Walkers, while Morning and Evening Walkers each comprised roughly 22% of the cohort.

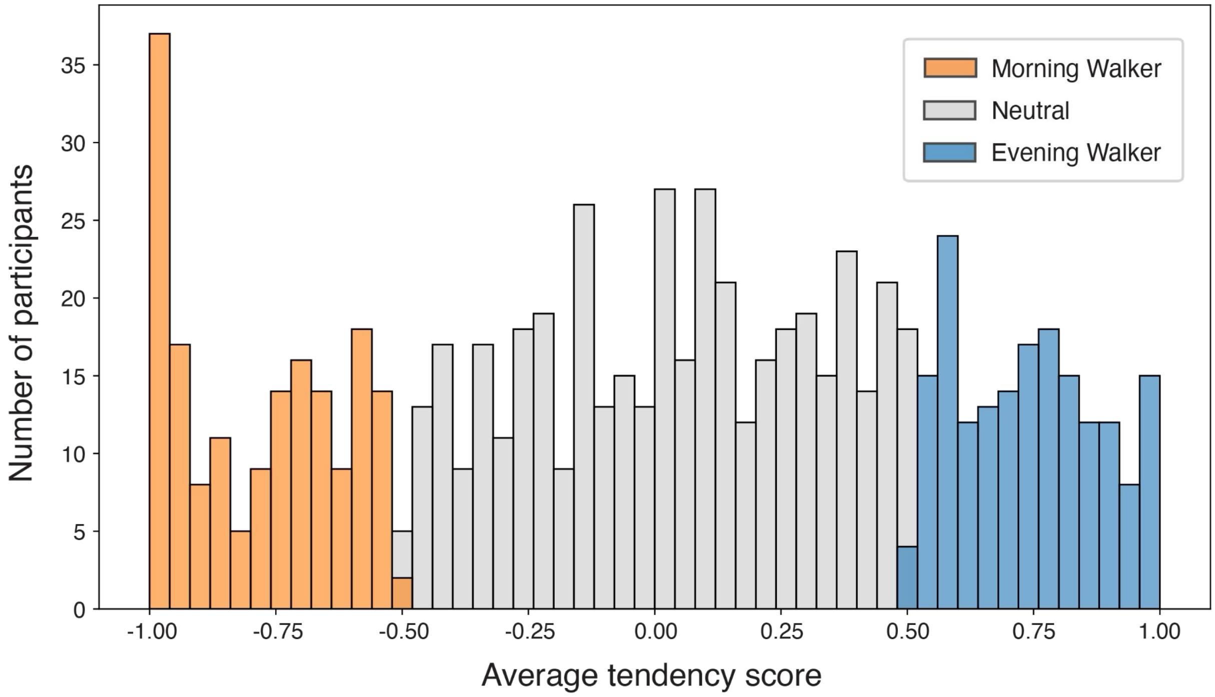
